# Health related quality of life and associated factors among postpartum women after normal vaginal delivery and caesarean section: A comparative cross-sectional study

**DOI:** 10.1101/2024.03.14.24304310

**Authors:** Wubet Mihretu Workneh, Simegnew Handebo, Temesgen Geleta, Biniam Yohannes Wotango, Bisrat Tamene Bekele

## Abstract

**Background:** Postpartum influences women’s physical health conditions and may affect their quality of life and future health. Some women suffer from health problems which are largely related to delivery mode. Assessing the postnatal quality of life is essential to address the concern of women and provide comprehensive postnatal care and minimizes the morbidity of the mother during and beyond the postnatal period.

**Objectives:** To assess health related quality of life and associated factors among postpartum women after normal vaginal delivery and caesarean section in public hospitals, Addis Ababa, Ethiopia.

**Methods:** A hospital-based comparative cross-sectional study was conducted in public hospitals of Addis Ababa using Short Form-36. Systematic random sampling method was used and telephone interviews were conducted for 171 Caesarian Section and 165 Normal Vaginal Delivery postpartum women. The collected data entered into Epi-info version 7.2 and then analysied by SPSS version 25. The Multiple linear regression model was used after checking the assumptions were met to identify associated factors and p-value of 0.05 and a confidence interval of 95% was employed.

**Result:** Overall 336 postpartum women participated with a response rate of 88%. The mean health-related quality of life was 91.42 and 81.89 for normal vaginal delivery and cesarean section group respectively.In multivariable linear regression for both groups, age =(β=-0.166;95%CI:-0.504,-0.121), family income (β=-0.188 95%CI-0.001,0.000), gestational age (β =0.131,95%CI:0.44,0.185), normal vaginal delivery (β=0.526; 95% CI: 6.790,9.621), and live birth (β=2.471,95% CI:1.094,9.643) were significantly associated factors and explained 47% of the variance in Health Related Quality of Life among postpartum women.

**Conclusion and Recommendation:** The study finds a higher level of health-related quality of life in the normal vaginal delivery group compared to the cesarean section. Health workers should give appropriate counseling on modes of delivery, inorder to help patients to make an informed decision about their childbirth experience.

## Introduction

Each year, about 210 million women become pregnant and about 140 million newborn babies are delivered—the sheer scale of maternal health alone makes maternal well being and survival vital concerns. Maternal health is key to sustainable development and to future generations. Poor maternal health as measured by mortality, morbidity and in recent times with quality of life during and beyond the postpartum period (1).

The postpartum or puerperium period, covers a critical transitional time for a woman, the newborn and the family as a whole during this period, a woman is adapting to multiple physical, social, and psychological changes. Recovering from childbirth, adjusting to changing hormones, and learning to feed and care for her newborn (2,3). In addition to being a time of joy and excitement, this “fourth trimester” can present considerable challenges for women, including lack of sleep, fatigue, pain, breastfeeding difficulties, stress and new onset or exacerbation of mental health disorders (4). This period starts about an hour after the delivery of the placenta and traditionally includes the following six weeks but up to six months considers as the delayed postpartum period (2,5).

Health related quality of life (HRQoL) for maternity populations can be defined as a multidimensional concept referring to a woman’s perception of the influence of her pregnancy, birth and postpartum condition, her care provision and any intervention and treatment on her physical, mental, emotional, and social functioning (7).

During the postpartum period and following childbirth, many women experience physical, emotional, functional, social and psychological challenges that can affect their well-being (10). In one study done in the USA, women 9–12 months post-partum found that 69% of women experienced at least one physical symptom following childbirth, while another study conducted in Spain reported that 85% of women experienced fatigue six weeks after childbirth (10,11). Some of these challenges are known to persist well beyond the traditional 6-week postpartum period and are associated with poor overall quality of life (QoL) among postpartum women (12).

Although the traditionally used pregnancy and postpartum period outcome measures, such as pregnancy-related morbidity and mortality rates, remain essential, they are no longer adequate on their own because population health should be assessed, not only on the basis of saving lives but also in terms of improving quality of life (14).

The perceived level of HRQoL plays a considerable role in an individual’s life since it describes the mental and physical health self-report. The pregnancy, birth and postpartum period, and a child being born along with the new role of the mother and the responsibilities that this entails, are significant periods in women’s lives that entail major changes in their quality of life due to physical, psychic and social repercussions (11). Maternal postpartum functioning plays an important role in maternal well-being as the postpartum year is a time of increased vulnerability to physical health concerns and is also the time period in which mothers are most likely to develop depression and anxiety compared to any other time in their life (10).

However, in Ethiopia and also in Africa little is known in this area and those studies show postnatal women experienced a significant impact on their health-related quality of life even though during the postpartum period women are experiencing fragmented maternity care.

### General Objective

The aim of this study is to assess Health related quality of life and associated factors among postpartum women after normal vaginal delivery and caesarean section in public hospitals, Addis Ababa, Ethiopia.

### Specific Objectives

- To compare the level of health related quality of life among postpartum women after normal vaginal delivery with caesarean section in public hospitals in Addis Ababa, Ethiopia.
- To identify the associated factors of health related qualities of life among postpartum women after CS and NVD in public hospitals in Addis Ababa, Ethiopia.

## Method and material

### Study area and period

Three public hospitals under AACAHB with high delivery load are selected. The first hospital is Gandhi Memorial Hospital which gives a maternal and neonatal related heath care service to the residents of Addis Ababa and surrounding community with up to 10,000 deliveries per year. Zewditu Memorial Hospital is known for its treatment of ART patients and currently treats over 6,000 each month, in its general health care service obstetric care is also delivered to the catchment population. Dagmawi Menillik II Hospital is a specialized hospital in ophthalmologic services and also different specialty care in addition to obstetrics care services.

The study was done from October 8, 2022 to February 6, 2023.

### Study Design

A hospital-based cross-sectional study was conducted.

### Source population

All women who gave birth in public hospitals and were within six months of after normal vaginal delivery and caesarian section in Addis Ababa were the source population

### Study Population

Selected postpartum women within six months after normal vaginal delivery and caesarian section at selected public hospitals were the study population.

### Inclusion criteria

Women within six months of normal vaginal delivery and caesarian section were included in this study.

### Exclusion criteria

Women having physical problems (spinal cord injury, amputation, paralysis, and deformity of the limb)

### Sample size determination

Sample size was estimated using sample size calculation formula for comparison between two mean for equal sample sizes. Sample size was calculated for different domains of SF-36 questioner which are mental health domain, social function domain and physical limitation domain. The largest sample size (382) was taken for which was determined using mental health domain. See *Table 1*

**Table 1.**
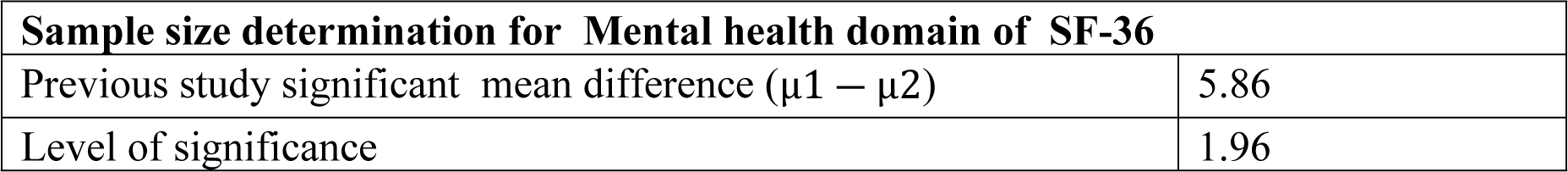
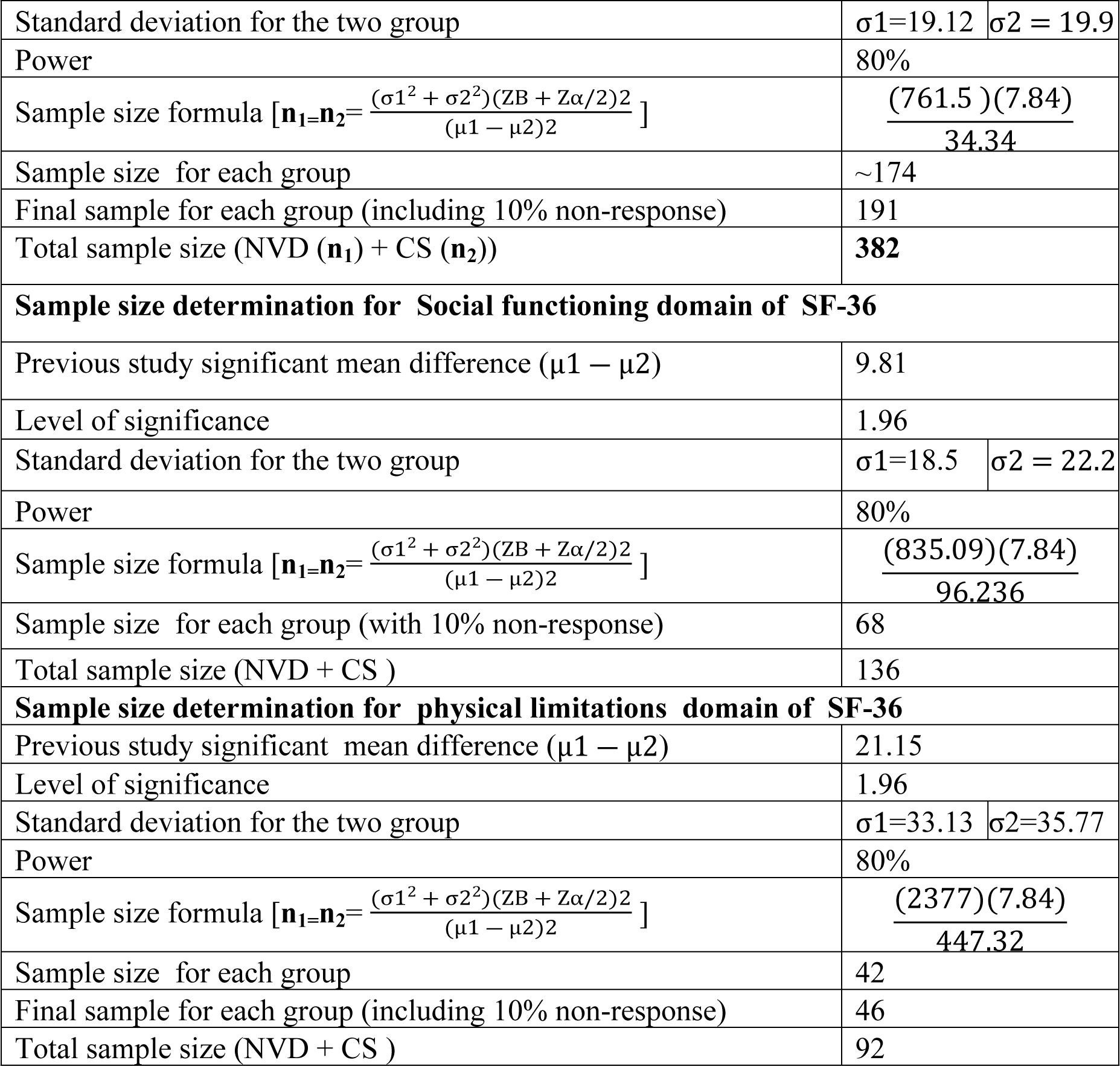
Sample size determination using significant mean of QoL difference between postpartum women giving birth by NVD and CS.

| **Sample size determination for Mental health domain of SF-36** |  |
| --- | --- |
| Previous study significant mean difference ($\mu_1 - \mu_2$) | 5.86 |
| Level of significance | 1.96 |

|  |  |  |
| --- | --- | --- |
| Standard deviation for the two group | $\sigma_1=19.12$ | $\sigma_2 = 19.9$ |
| Power | 80% |  |
| Sample size formula $[n_1=n_2=\frac{(\sigma_1^2 + \sigma_2^2)(ZB + Z\alpha/2)^2}{(\mu_1 - \mu_2)^2}]$ | $\frac{(761.5)(7.84)}{34.34}$ | |
| Sample size for each group | ~174 |  |
| Final sample for each group (including 10% non-response) | 191 |  |
| Total sample size (NVD ($n_1$) + CS ($n_2$)) | 382 | |
| Sample size determination for Social functioning domain of SF-36 |  |  |
| Previous study significant mean difference ($\mu_1 - \mu_2$) | 9.81 | |
| Level of significance | 1.96 |  |
| Standard deviation for the two group | $\sigma_1=18.5$ | $\sigma_2 = 22.2$ |
| Power | 80% |  |
| Sample size formula $[n_1=n_2=\frac{(\sigma_1^2 + \sigma_2^2)(ZB + Z\alpha/2)^2}{(\mu_1 - \mu_2)^2}]$ | $\frac{(835.09)(7.84)}{96.236}$ | |
| Sample size for each group (with 10% non-response) | 68 |  |
| Total sample size (NVD + CS ) | 136 |  |
| Sample size determination for physical limitations domain of SF-36 |  |  |
| Previous study significant mean difference ($\mu_1 - \mu_2$) | 21.15 | |
| Level of significance | 1.96 |  |
| Standard deviation for the two group | $\sigma_1=33.13$ | $\sigma_2=35.77$ |
| Power | 80% |  |
| Sample size formula $[n_1=n_2=\frac{(\sigma_1^2 + \sigma_2^2)(ZB + Z\alpha/2)^2}{(\mu_1 - \mu_2)^2}]$ | $\frac{(2377)(7.84)}{447.32}$ | |
| Sample size for each group | 42 |  |
| Final sample for each group (including 10% non-response) | 46 |  |
| Total sample size (NVD + CS ) | 92 |  |

### Sampling Technique and procedures

A total of 7808 deliveries by NVD and CS was attended in selected hospitals within a six moths period.

After counting both the normal vaginal delivery and cesarian section separately of six months from the delivery room HMIS register, the sample was allocated for each hospital using proportionate to the size.

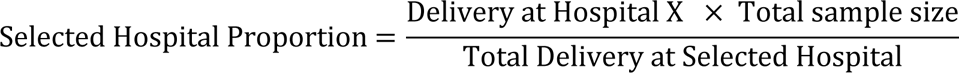

A systematic random sampling technique was used for sample selection. For GMH the 2090 NVD and 1710 CS registered were divided by the sample size (93) and we get 22 and 18 respectively then the first case was determined by the lottery method after that the rest of the sample was taken every 22^nd^ and 18^th^ and recorded to the abstraction format prepared by the researcher. The phone number was obtained from the patient’s chart guided by the patient’s name and medical record number. In case of incomplete documentation and unclear handwriting, the next immediate case was taken. For ZMH the 964 CS and 1386 NVD cases were devided by 57 and 17 and 24 were the respected numbers then after determining the first cases by the lottery method the rest of the sample was taken every 17^th^ and 24^th^ for CS and NVD cases respectively. For DMIIH by employing the same procedure the first case was determined using lottery method every 15^th^ the CS was selected and every 29^th^ the NVD cases selected.

### Operational definitions

- **Health related quality of life (HRQoL)** Health-related quality of life indicates the extent to which a disease or medical condition impacts upon the daily physical, emotional, mental and contextual well-being of an individual. It was measured by SF-36 tool which comprises eight scaled scores.The items are responded through 2,3,5 and 6-option Likert scale and a score between 0 and 100 is assigned to each dimension. The scores are weighted sums of the questions in each section. Scores range from 0 - 100 Lower scores = more disability, higher scores = less disability.
- **Normal Vaginal Delivery** – Vaginal delivery with natural process that usually does not require significant medical intervention with or without episiotomy procedures.
- **Caesarian Section** - a surgical operation for delivering a child by cutting through the wall of the mother’s abdomen.
- **Postpartum period**- begins immediately after childbirth up to six months.

### Data collection tool

The data were collected using semi-structured questionnaires adapted from different peer-reviewed published studies (10,14,15,19). The questionnaire is composed of three parts socio-demographic characteristics of the mother, obstetric and medical history and neonatal-related questionnaire, and health-related quality of life questionnaire. A structured pre-tested interviewer-administered questionnaire was developed in English and then translated into the Amharic language for simplicity and then back translated to the English language for its consistency by two different individuals who speak both English and Amharic fluently.

Standardised MOS SF-36 tool which was validated and translated for Ethiopia by a validation study was used to assess HRQoL. The tool was validated for self-administration and for administration by a trained interviewer in person or by telephone. According to a study done in Butajira, rural Ethiopia, the SF-36 appears to be an appropriate measure for measuring health related quality of life in various population groups in Ethiopia with confirmed validity of the questionnaire with Cronbach’s alpha higher than 0.70 for physical functioning, role limitations due to physical health, bodily pain, general health, emotional well being, role limitations due to emotional problems, vitality except for social functioning which was 0.68 (23).

### Data collection procedures

The data collection for selected respondents was done through telephone interview within six months of their postpartum period. The Amharic version of the structured questionnaire was printed in paper. Completed questionnaires were collected and given to the principal investigator on the day of data collection. In cases where respondents’ couldn’t answer their phone or their phone was not working, the data collectors tries on another time and in order to declare as nonrespondents five phone calls must be attemted. Each data collector did have a code to make data management easy. The data collection was conducted by four Midwives who hold BSc degrees and was supervised and closly followed by the principal investigator.

### Data quality assurance

The quality of data was controlled starting from the time of questionnair preparations. The questionnaire was selected by reviewing relevant literatures on the subject to ensure reliability (23,24). The Amharic version of the SF-36 questioner was used (25). Training was conducted for data collectors on the purpose of study, procedures of data collection for one day prior to the study. There was periodic supervision and technical support for data collectors. After completing the training, trainees conducted pre-test using 5% of the total sample size which was 16 at non-study heath facility which was RDDH. The data collection was conducted by 4 midwives and supervised by the principal investigator. In addition to this, a close follow-up by the principal investigator was done. During data collection, the supervisor received questionnaires from data collectors and review for completeness, accuracy, and consistency on daily bases. Incomplete, inconsistent and invalid data was refined properly to get maximum quality of data before, during and after data entry.

### Data processing and analysis

After data collection, each questionnaire was checked for completeness based on the code given during data collection. Data was entered in to Epi Info Version 7.2 then was exported to Statistical Package for Social Studies (SPSS) version 25. Coding of individual questionnaires was checked before data entry in to the software. Further, data cleansing was performed to check for outliers, missed values and any inconsistencies before the data was analyzed using the software. Descriptive data analysis was performed using means and medians and continuous variables was be expressed in mean and standard deviation and mean score of health related quality of life was compared using independent group t-test.

The health related quality of life score were approximately normally distributed, so untransformed score was used for the bivariate and multivariable regression analysis. Multiple linear regressions was used to explore the real difference in quality of life score and to identify significant predictor factor of health related quality of life of each study group. Only variables that were statistically significant at a p-values of less than 0.2 in the bivariate analysis were included in the multivariable models. Scatter plot was used to check the linearity, box plots were used to identify potential outliers. Variance inflation factors (VIF) used to diagnose the multicollinearity and equality of variance was checked using Levene’s test to check if the standard deviation of the residual errors is equal across all independent variables.Models were tested for model fit. In performing the statistical analyses p value of 0.05 and confidence interval (CI) of 95% was employed. To control the effect of confounding variables stepwise multiple linear regression analysis was done.

## RESULTS

A total of 336 participants were included in this study (giving an 88 % response rate). Fourty-six women were not included, thirty-nine of them due to being unable to reach their phone, while seven of them rejected their participation in the study. Of the total respondents, 171 were delivered via CS while 165 were by NVD.

### Sociodemographic Characteristics of the study participants

The mean age of women was 26.22 ± 3.693 standard deviations (SD) and 27.73 ± 4.422 standard deviations (SD) for normal vaginal birth and caesarean group, respectively. Nearly 90% (146) of the normal vaginal birth group lived in urban areas compared to 139 (81.3%) of the caesarean group. Regarding the level of education, 76 (46.1%) of NVD and 83 (48.5%) of CS groups were completed secondary education. Moreover, 68 (39.8%) of CS group were private employee while 58 (35.2%) of NVD were housewives. The mean monthly family income was 6503.64 ± 3231.870 birr and 6923.98 ± 3130.506 birr for NVD and CS group, respectively. About three hundred twenty nine participants answered their husbands’ occupation and majority accounts for private job 103 (60.3%) in CS group and 83 (50.3%) NVD group. See *Table 2*

**Table 2.** Sociodemographic characteristics of the study participants at selected public hospitals in Addis Ababa, Ethiopia.

| Variables | Response | Caesarian Section (N=171) |  | Normal vaginal Delivery (N=165) |  |
| --- | --- | --- | --- | --- | --- |
|  |  | Frequency | Percent | Frequency | Percent |
| Age in years | 18-24 | 40 | 23.4% | 56 | 33.9% |
|  | 25-34 | 118 | 69.0% | 102 | 61.8% |
|  | 35-45 | 13 | 7.6% | 7 | 4.2% |
| | Mean ± SD | 27.73 ± 4.422 | | 26.22 ± 3.693 | |
| Residence | Urban | 139 | 81.3% | 146 | 88.5% |
|  | Rural | 32 | 18.7% | 19 | 11.5% |
| Marital status | Married | 167 | 97.7% | 153 | 92.7% |
|  | Single | 4 | 2.3% | 12 | 7.3% |
| Level of education | Read and Write | 14 | 8.1% | 6 | 3.6% |
|  | Primary education (1-8) | 44 | 25.7% | 44 | 26.7% |
|  | Secondary education (9-12) | 83 | 48.5% | 76 | 46.1% |
|  | College/University | 30 | 17.5% | 39 | 23.6% |
| Occupation | House wife | 45 | 26.3% | 58 | 35.2% |
|  | Daily laborer | 7 | 4.1% | 14 | 8.5% |
|  | Private Employee | 68 | 39.8% | 42 | 25.5% |
|  | Government employee | 29 | 17.0% | 30 | 18.2% |
|  | Private Job | 22 | 12.9% | 21 | 12.7% |
| Monthly family income (Ethiopian Birr) | Mean ± SD | 6923.98 ± 3130.506 | | 6503.64 ± 3231.870 | |
| Husband's Occupation | Private | 103 | 60.3% | 83 | 50.3% |
|  | Daily Laborer | 26 | 15.2% | 14 | 8.5% |
|  | Government Employee | 24 | 14.0% | 37 | 22.4% |
|  | NGO | 13 | 7.6% | 19 | 11.5% |

N stands for frequency while SD is for standard deviation

### Obstetric and health related factors

Of the total CS births, 88(51.5%) mothers were multipara while 117 (70.9%) of NVD were primipara. From the total of 136 multiparous mothers of both groups, 88 (64.7%) gave birth via CS. See *Table 3* About 336 (98.2%) mothers had ANC follow-up. The mean number of ANC visit was 7.56 ± 2.440 and 5.95 ± 2.261 for CS and NVD respectively.

**Table 3.** Obstetrics and health related factors of the study participants at selected pubic hospital in Addis Ababa, Ethiopia.

| Variables | Response | Caesarian Section (N=171) |  | Normal vaginal Delivery (N=165) |  |
| --- | --- | --- | --- | --- | --- |
|  |  | Frequency | Percent | Frequency | Percent |
| Parity | Primipara | 83 | 48.5% | 117 | 70.9% |
|  | Multipara | 88 | 51.5% | 48 | 29.1% |
| ANC follow up | Had ANC | 169 | 98.8% | 161 | 97.6% |
|  | No ANC | 2 | 1.2% | 4 | 2.4% |
| Number of ANC | Mean ± SD | 7.56 ± 2.440 | | 5.95 ± 2.261 | |
| Obstetric Complication | Yes | 49 | 28.7% | 37 | 22.4% |
|  | No | 122 | 71.3% | 128 | 77.6% |
| Obstetric Complication | APH | 12 | 24.5% | 4 | 10.8% |
|  | PROM | 10 | 20.4% | 26 | 70.3% |
|  | PIH | 27 | 55.1% | 7 | 18.9% |
| Gestational Age (in weeks) | Median | 39.0 |  | 40.2 |  |
| Medical Illness | Yes | 10 | 5.8% | 4 | 2.4% |
|  | No | 161 | 94.2% | 161 | 97.6% |
| Medical Illness by Type | Hypertension | 6 | 60% | 0 | 0% |
|  | Diabetes | 0 | 0% | 4 | 100% |
|  | Epilepsy | 4 | 40% | 0 | 0% |

N stands for frequency while SD is for standard deviation

Regarding obstetric complications, 49 (28.7%) CS group and 37 (22.4%) NVD group had a complication. The majority of the NVD group 26 (70.3%) and 27 (55.1%) of the CS group reported premature rupture of membranes and pregnancy induced hypertension respectively as a complication. The median gestational age for CS and NVD groups were 39 weeks and 40 weeks respectively. About 14 participants from both groups responded to existing medical illnesses during their postpartum period which were six hypertensive, four diabetes and four epilepsy.

### Newborn related factors

About 176 (52.4%) of neonates born from both groups were female while 160 (47.6%) were male. Concerning neonatal delivery outcome status, 167(97.7%) Caesarean birth and 162 (98.2%) normal vaginal birth were live births. About 22 (12.9%) of neonates from CS group and 14 (8.5%) of NVD were hospitalized.

### Comparison of magnitude of Health Related Quality of Life of Study participants

Test of normality was done for HRQoL, physical component summary (PCS) and mental component summary (MCS) and untransformed values were analysed using an independent sample t-test.

There were statistically significant differences between CS and NVD groups in total HRQoL, PCS and MCS. From the eight domains of the SF-36 questionnaire, five of the domains with statistical difference were physical functioning, role limitations due to physical health, bodily pain, general health and vitality. See *Table 4*.

**Table 4.** Comparison of magnitude of Health Related Quality of Life of Study participants in selected public Hospitals of Addis Ababa, Ethiopia.

| SF-36 HRQoL |  | NVD<br>(N=165)<br>Mean ± SD | CS<br>(N=171)<br>Mean ± SD | Mean<br>difference | 95% CI |  | P-value |
| --- | --- | --- | --- | --- | --- | --- | --- |
|  |  |  |  |  | Lower | Upper |  |
| PCS | Number<br>of items |  |  |  |  |  |  |
| Physical<br>functioning | 10 | 93.60±10.62 | 75.65±10.61 | 17.99 | 15.71 | 20.26 | < 0.001* |
| Role limitations<br>due to physical<br>health | 4 | 91.96±19.12 | 71.34±26.24 | 20.62 | 15.70 | 25.54 | < 0.001* |
| Bodily Pain | 2 | 90.32±17.19 | 64.49±17.54 | 25.82 | 22.09 | 29.55 | <0.001* |
| General health | 5 | 88.04±9.47 | 78.53±10.09 | 9.50 | 7.40 | 11.60 | <0.001* |
| Total PCS |  | 90.98±10.38 | 72.49±12.12 | 18.48 | 16.06 | 20.90 | < 0.001* |
| MCS |  |  |  |  |  |  |  |
| Social<br>Functioning | 2 | 98.56±4.00 | 98.46±4.11 | 0.09 | -0.77 | 0.96 | 0.829 |
| Emotional well<br>being | 5 | 82.98±4.70 | 82.19± 3.73 | 0.78 | -0.13 | 1.69 | 0.093 |
| Role limitations<br>due to emotional<br>problems | 3 | 98.18±9.20 | 99.02 ±5.63 | -0.84 | -2.48 | 0.80 | 0.314 |
| Vitality | 4 | 87.73±3.44 | 85.46±4.32 | 2.25 | 1.41 | 3.10 | <0.001* |
| Total MCS |  | 91.86±3.03 | 91.28±2.16 | 18.48 | 16.06 | 20.90 | 0.048* |
| **HRQoL** |  | 91.42±5.97 | 81.89±6.40 | 9.52 | 8.20 | 10.85 | < 0.001* |

N stands for frequency while SD is for standard deviation, \* Statistically significant = $p \leq 0.05$ , PCS=physical component summary, MCS=Mental component summary

### Factors associated with HRQoL

Simple and multivariable linear regression was done to display the significance of each independent variable in predicting the HRQoL after checking the assumptions of linear regression analysis.

The socio-demographic variables, obstetrics and medical health related and neonatal related characteristics of postpartum women who gave birth by NVD and CS at public hospitals were analyzed in bivariate analysis then variables that were significant at different degree of P-value ≤ 0.05 included in multivariate analysis.

Three regression models were used to compare the NVD and CS groups of HRQoL. The first model was used to identify the predictors of HRQoL in the general population, age, marital status, family income, parity, number of ANC, gestational age, medical illness type, mode of delivery and delivery outcome which were significant in the simple regression model were included. The second regression model was done for the CS groups, factors such as age, resident, marital status, family income, parity, number of ANC, sex of newborn, and delivery outcome which was significant in a simple regression model were computed. And Age, gestational age, previous mode of delivery and delivery outcome were included in the NVD group.

While doing the analysis standardized beta coefficient was used to compare the strength of the effect of each individual independent variable to the dependent variable.

### Multiple linear regression analysis: Factors associated with the componenent of HRQoL in all Population

Age, family income, gestational age at the time of delivery, number of ANC visit, mode of delivery and delivery outcome were significantly associated factors with total HRQoL with, F(15,320)=31.669, P<0.001 and adjusted R^2^ = 48.2%. See *Table 5*.

**Table 5.** Summary of multiple linear regession on HRQoL with independent variables of all population in public hospitals, Addis Ababa, Ethiopia.

| Variable | Categories | HRQoL |  |  |
| --- | --- | --- | --- | --- |
| | | B | $\beta$ (95% CI) | P value |
| Age |  | -0.312 | -0.166 (-0.504,-0.121) | 0.001* |
| Marital Status | Single(Ref) |  |  |  |
|  | Married | -1.237 | -0.034(-4.300,1.827) | 0.428 |
| Family Income |  | -0.000 | -0.188 (-0.001,0.000 ) | 0.007* |
| Occupation | House wife(ref) |  |  |  |
|  | Daily Laborer | 1.101 | 0.034(-1.472,3.674) | 0.400 |
|  | Private Employee |  |  |  |
|  | Governement Employee |  |  |  |
|  | Private Job |  |  |  |
| Husband Occupation | Private (Ref) |  |  |  |
|  | Daily Laborar | 2.046 | 0.085(0.024,4.067) | 0.057 |
|  | Governement Employee |  |  |  |
|  | NGO |  |  |  |
| Parity | Multipara(Ref) |  |  |  |
|  | Primipara | 1.016 | 0.064(-0.575,2.608) | 0.210 |
| ANC Followup | No(Ref) |  |  |  |
|  | Yes | 1.827 | 0.031(-3.217,6.870) | 0.477 |
| Number of ANC |  | -0.382 | -0.122(-0.676,-0.88) | 0.011* |
| Gestational Age |  | 0.114 | 0.131 (0.44,0.185) | 0.002* |
| Type of Obsatrics Complication | APH(Ref) |  |  |  |
|  | PROM | -0.754 | -0.030(-2.851,1.343) | 0.480 |
|  | PIH | 1.232 | 0.048((-0.876,3.339) | 0.251 |
| Medical Illness | No(Ref) |  |  |  |
|  | Yes | 8.170 | 0.209(4.451,11.889) | 0.080 |
| Medical Illness Type | Hypertension(Ref) |  |  |  |
|  | Diabetes | -0.061 | -0.001(-6.835,6.714) | 0.986 |
|  | Epilepsy |  |  |  |
| Mode of Delivery | CS(Ref) |  |  |  |
|  | NVD | 8.206 | 0.526 (6.790,9.621) | <0.001* |
| Delivery Outcome | Stillbirth(Ref) |  |  |  |
|  | Alive | 5.369 | 2.471(1.094,9.643) | 0.014* |

For every one year increase in age of postpartum women and for every one unit increase in total family income HRQoL decreases by 0.166 and 0.188 times respectively (β = - 0.166, t= −3.206, p = 0.001and β = −0.188, t= −2.698, p = 0.007).

For one visit increase in a ANC follow up HRQoL decreases by 0.122 (β = −0.122, t= −2.557 p=0.011)

Postpartum women who gave birth by NVD had 0.526 times more HRQoL compared to those who gave birth by CS (β = 0.526, t=11.405, p <0.001). For every one week increase in gestational age at the time of delivery also increases HRQoL by 0.131 (β = 0.131, t=3.182, P=0.002).

A multivariate analysis of delivery outcome indicates those postpartum women who gave birth to alive neonates had 0.098 times more HRQoL compared to those who gave birth to stillborn (β = 0.098, t=2.471 p =0.014).

## Disscussion

This study found significant differences in the level of individual domains of the SF-36 tool and HRQoL between NVD and CS groups.

The results of this study showed that the health-related quality of life during the postpartum period was higher in the NVD group compared to the CS group which is consistent with the study done in India and Egypt, and Nigeria (10,19,26). The results of this study demonstrate that if given the choice of birth method, women may want to consider a natural vaginal delivery due to the association with a better health-related quality of life during the postpartum period. So that women should be informed about all the potential benefits and risks associated with both types of birth prior to making an informed decision about their delivery method. Whereas the study done in china and Iran showed no significant difference between the two groups (11,12), in the study done in Ethiopia cesarean delivery identified to have a positive association with better HRQoL (15).

In all the physical component summary domains which are physical functioning, role limitations due to physical health, bodily pain, and general health NVD groups had superior mean scores compared to the CS groups. This finding is agreeing with the Iranian study and was partly comparable to the Egyptian study which reported the same finding except in the general health domain (13,19). The explanation for the differences in the NVD and CS groups might due to the surgical procedure, and the effect of anesthesia undergone in CS groups further affect their postpartum recovery compaired to vaginal birth. The surgical procedure and the medical intervention might cause delayed postpartum recovery and low quality of life. This finding has important implications for both medical practitioners and patients.

From a medical perspective, it is important to recognize that vaginal delivery is associated with superior physical health outcomes compared to cesarean sections. In particular, vaginal delivery is associated with better physical functioning, less role limitation due to physical health issues, reduced bodily pain, and improved overall general health. This suggests that health professionals should prioritize vaginal delivery when possible so that patients can benefit from these improved physical health outcomes.

From a patient perspective, this finding highlights the importance of considering all available information when making choices about childbirth. While Cesarean sections are often necessary and beneficial in certain circumstances, this research suggests that if a pregnancy allows for it, vaginal delivery should be the preferred choice for improved physical health outcomes in the future. By choosing vaginal delivery whenever possible, patients can significantly improve their long-term physical wellbeing.

Mental component summaries of the SF-36 and the vitality domain had significantly higher scores in NVD compared to the CS group which was consistent with the study done in Iran, This might be because of the emotional strains that the surgery imposes on a woman’s body (13).

In a multivariate analysis exploring the regression model, older age, monthly income of a family and number of ANC visits were factors that are associated with lower HRQoL in all populations of the respondents while increased gestational age during delivery, giving birth by NVD and havening alive neonate were the factors that are associated with better HRQoL. For CS groups monthly family income and having pregnancy-induced hypertension as an obstetric complication compared to the other complications were associated with lower postpartum HRQoL while having a male sex child was associated with increased HRQoL. Delivering an alive neonate and increased gestational age at the time of delivery were factors that increased the HRQoL of postpartum women in the NVD group while being older women associated with decreased HRQoL.

The same as the study done in Arbaminch, Ethiopia and Spain my study found that when the age of women and postpartum HRQoL had a negative association (15,17). This association might be due to the fact that when a woman’s age increases the probability of having both medical and obstetrics complications increases and this might compromised the quality of life. As women age, they tend to experience an increased risk of developing certain chronic illnesses that can lead to physical and emotional complications. Additionally, age can be associated with inflammation in the body, fatigue and muscular weakness which could affect mental health and well-being. These changes could result in a lower health-related quality of life, especially during the postpartum period.

Healthcare providers should consider the potential effects of a woman’s age on her health-related quality of life during the postpartum period. When assessing the long-term needs of postpartum women after delivery, healthcare workers should take into account the increased risks that come along with advancing age. Healthy lifestyle practices such as regular exercise, adequate sleep hygiene and proper nutrition should be discussed with older mothers so as to ensure improved physical and mental wellbeing for them during this vulnerable period.

Similarly, the monthly income of the family had a negative association with HRQoL this finding is in contrast with the Iran study which found that income is an important factor for better QoL(12). The finding of my study might be attributed to women who had high economic status might have higher expectations and perceptions of quality of life. In addition, higher income can be associated with decreased well-being, such as a decrease in overall life satisfaction amongst affluent individuals. Therefore, the finding suggests that even though women may obtain financial security and other resources through an increase in income after childbirth, they may not experience improved quality of life or well-being as a result.

The gestational age at which women gave birth was also identified as a risk factor for a worse HRQoL score in all respondant populations and in women who gave birth by NVD in these study, which conforms with other studies (17). This is because preterm deliveries might associate with a bad neonatal outcome like hospitalization and stillborn and due to this mothers might encounter emotional and social problems. This finding reinforces the previous body of evidence that suggests greater gestational age is associated with improved health outcomes for infants and women during pregnancy and childbirth.

These study found that women who gave birth alive child had higher HRQoL compared to postpartum women who had stillborn, this implies that pregnant women and their neonates are both receiving improved healthcare outcomes. Furthermore, the research indicates that having a live infant leads to an overall improvement in the quality of life for mothers during the postpartum period. This suggests that with proper antenatal care and support services, mothers can have a better physical and mental health status during this time. Additionally, it shows the importance of providing suitable resources for mothers so they can provide their neonates with proper care during the delivery process and afterwards. The findings justify why it is essential for healthcare providers to invest in improving maternal healthcare access and outcomes for pregnant women before, during and after pregnancy in order to facilitate better maternal physical and mental well-being later on in life.

Postpartum women who gave birth via CS and had male neonates had higher HRQoL compared to women with female neonates this is inconsistence with finding in China community the difference might be attributed to the desire and positive influence of having male neonates culturally in the Ethiopian community (11).

## Limitations of the Study

This research is not without limitations, the first limitation is that the study was only done in the postpartum period and did not include any time before birth. There could be recall bias on certain variables during a phonecall interview. The other limitation of this study is quality-of-life determining factors like behavioral risk, depression, other psychiatric states, and health service related factors are not included in this study.

## Conclusion and Recommendations

This study has demonstrated that women who undergo vaginal delivery, compared to those who deliver via Cesarean section, have markedly better physical and mental health outcomes in the postpartum period. Improved physical health outcomes include greater physical functioning, less role limitation due to physical health issues, less bodily pain, and improved general wellbeing. Furthermore, better mental health outcomes were indicated by higher scores in both the Mental Component Summary of the SF-36 and in the Vitality domain specifically. These results highlight the importance of considering all available information when making decisions about childbirth as opting for a vaginal delivery when possible could significantly improve a mother’s long-term physical and mental health outcomes. Therefore, health professionals should prioritize vaginal delivery when possible so that patients can benefit from these superior physical health outcomes and improved quality of life, while patients should be informed about all available information so they can make an informed decision about their childbirth experience.

## Data Availability

All relevant data are within the manuscript and its Supporting Information files.

## Abbreviations

AACAHB: Addis Ababa City Administration Health bureau
CS: Caesarian section
DRQoL: Disease specific health related quality of life
DMIIH: Dagmawi Menelik II Hospital
GCS: Generic Core Scale
GMH: Gandhi Memorial Hospital
HRQoL: Health Related Quality of Life
MCS: Mental Component Summary
MOS: Medical Outcomes Study
NVD: Normal vaginal delivery
PCS: Physical Component Summary
PPD: Postpartum Depression
QoL: Quality of Life
RDDH: Ras Desta Damittw Hospital
WHO: World Health Organization
ZMH: Zeweditu Memorial Hospital

## Authors contributions

All authors made a significant contribution to the study in the conception, study design, execution, acquisition of data, analysis and interpretation: involved in drafting, substantially revising and critically reviewing the article; have agreed on the journal to which the article will be submitted. All authors reviewed and agreed on the revised versions of the article before submission and agreed to take responsibility and be accountable for all aspects of the work.

## Availability of data and materials

All data generated or analyzed during this study are available from the corresponding author on reasonable request.

## Ethical considerations and Informed Consent

Ethical approval was sought from St. Paul’s Hospital Millennium Medical College, Institutional Research Review Board (IRB) with with Ref. No. of ሕ/ጤ/ት/822/14. Ethical clearance was obtained from Addis Ababa city administration health bureau research directorate with Ref. No. of A/A/554/227. Then, permission to carry out the study was sought from GMH, ZMH and DMIIH Hospitals’ administrations. Oral consent was obtained from all participants after explaining the aim of the study and confirmed that the information would be used for research purpose and they had the right to refuse at any time. Confidentiality of the information obtained from the interview was maintained throughout the process of data collection. This study was conducted as per the Declaration of Helsinki.

## Acknowledgements

The authors would like to acknowledge the study participants, data collectors, and administrative bodies of GMH, ZMH, DMIIH, and RDDH for their permission.

